# Heterologous vaccination as a strategy to minimize inequity in COVID-19 vaccine access: A modeling study in Thailand

**DOI:** 10.1101/2022.08.05.22278489

**Authors:** Suparinthon Anupong, Tanakorn Chantanasaro, Chaiwat Wilasang, Natcha C. Jitsuk, Chayanin Sararat, Kan Sornbundit, Busara Pattanasiri, Sudarat Chadsuthi, Charin Modchang

## Abstract

**Background:** Vaccinations are the best hope to control the COVID-19 pandemic and save lives. Due to the high demand and failure to share vaccines equitably, there were not enough vaccine supplies to cover the majority of people in low- and middle-income countries during the early stage of vaccination. To cope with this problem, Thailand, an upper-middle-income country, decided to employ a heterologous vaccination strategy as the primary COVID-19 vaccination regimen in the country. The CoronaVac (CV) vaccine was administered as the first dose, followed by the ChAdOx1 nCoV-19 (AZ) vaccine as the second dose. However, there is no study to assess the effectiveness of the heterologous vaccination employed in Thailand compared to the standard homologous vaccination.

**Methods:** We delineated the course and timeline of COVID-19 vaccination in Thailand. An age-structured compartmental model for COVID-19 transmission and vaccination was constructed and employed to assess the effectiveness of the heterologous vaccination strategy. The impact of the vaccine prioritization strategies on COVID-19 mortality and infections was also investigated.

**Results:** We found that the CV+AZ heterologous vaccination strategy outperforms the separate CV and AZ homologous vaccinations in reducing cumulative cases and deaths when combined with other non-pharmaceutical interventions. Furthermore, the results suggested that prioritizing vaccines for the elderly could be optimal in reducing COVID-19 mortality for a wide range of vaccination rates and disease transmission dynamics.

**Conclusions:** Our modeling results suggested that to minimize the impacts of inequity in early COVID-19 vaccine access in low- and middle-income countries, those countries may use early accessible but maybe lower-efficacy vaccines as the first dose of heterologous vaccination in combination with higher-efficacy vaccines as the second dose when they are available.

## Introduction

Since the coronavirus disease 2019 (COVID-19) was first detected in December 2019, it has spread to more than 100 countries [1], causing more than 5.4 million deaths and 286 million infected cases worldwide by the end of 2021 [2]. The pandemic is far from over, and vaccination, in combination with nonpharmaceutical interventions, is still the most reliable approach to prevent and mitigate the spread of the disease. However, although COVID-19 vaccines are effective in fighting against the disease [3-8], they are beneficial only when appropriately administered.

Although 58% of the world population received at least one dose of a COVID-19 vaccine at the end of 2021, there was still inequality in early vaccine accessibility. In particular, while over 70% of the population in wealthy nations completed the initial vaccination protocol at the end of 2021, only 4% of people in low-income countries have been vaccinated with at least one dose of COVID-19 vaccines [9]. In addition, the vaccines administered in high-income countries were mainly the mRNA vaccines, which have higher efficacy than the inactivated virus vaccines used in low- and middle-income countries [10, 11].

In Thailand, an upper-middle-income country, COVID-19 vaccines were first available in late February 2021 [9]. CoronaVac by Sinovac Biotech Ltd. (CV) and ChAdOx1 nCoV-19 (Vaxzevria) by AstraZeneca plc (AZ) vaccines were mostly deployed in the early stages of the vaccination campaign in Thailand. However, due to the emergence of the Delta variant and the shortage in AZ vaccine supply [12], the Department of Disease Control of Thailand decided to employ a heterologous vaccination strategy as the primary COVID-19 vaccination regimen in Thailand on July 12, 2021 [13]. For this regimen, the CV vaccine was administered as the first dose, followed by the AZ vaccine as the second dose. Although this CV-AZ heterologous vaccination strategy could help accelerate the vaccination speed in Thailand as the second dose could be administered 3 weeks after the first dose compared to 12 weeks in the traditional AZ-AZ homologous vaccination, there is still no study to assess the effectiveness of this CV-AZ heterologous vaccination over implementing the standard separate CV-CV and AZ-AZ homologous vaccination regimens. If this CV-AZ heterologous vaccination is proven to be superior to the corresponding homologous vaccination strategies, it could serve as a lesson learned for the low- and middle-income countries for combating a future emerging disease by using an early accessible (but maybe with lower efficacy) vaccine as the first dose, and then following by a higher efficacy vaccine as a booster or a second dose when they are available.

In this study, we, therefore, aimed to assess the effectiveness of the CV-AZ heterologous vaccination strategy using Thailand as a case study. Specifically, we first delineated the course and timeline of COVID-19 transmission and vaccination in Thailand. An age-structured compartmental model for COVID-19 transmission was then employed to assess the effectiveness of the implemented heterologous vaccination strategy. Finally, the impact of vaccine prioritization strategies was also investigated.

## Materials and methods

### Data sources

The numbers of confirmed COVID-19 cases and deaths in Thailand were retrieved from the Department of Disease Control, Ministry of Public Health, Thailand [14, 15]. Data on vaccine administration, including the numbers of the first, second, and booster doses, were obtained from refs. [16, 17]. The data on administered vaccine doses by the manufacturers were collected from ref. [16].

### COVID-19 transmission and vaccination model

We developed an age-structure compartmental model with two-dose vaccination to analyze the dynamics of COVID-19 transmission in Thailand. The schematic of the model is shown in **Figure 1**. Based on the available contact data in Thailand [18], the population was classified into 16 age groups with a five-year interval (0-4, 5-9, …, 70-74, and ≥ 75 years). All individuals were assumed to be initially susceptible (S) to SARS-CoV-2 infection. When susceptible individuals are infected, with the force of infection *λ*, they immediately transition to the exposed compartment (E). The exposed individuals then move to the infectious compartment with a rate *σ* that is inversely proportional to the latent period. The infectious individuals can be either symptomatic (I) or asymptomatic (A). The proportion of asymptomatic infections (*fA*) was assumed to be the same for all age groups. A fraction of *fSD* of symptomatic infectious individuals dies with a rate of *τ* that is inversely proportional to the duration from symptomatic infectious to death. The age-specific *fSD* was estimated from the age-specific infection fatality rate (*IFR*) obtained from refs. [19, 20]. Both I and A move to the recovered compartment RS and RA, respectively, with a rate *γ* that is inversely proportional to the infectious period.

**Figure 1.**
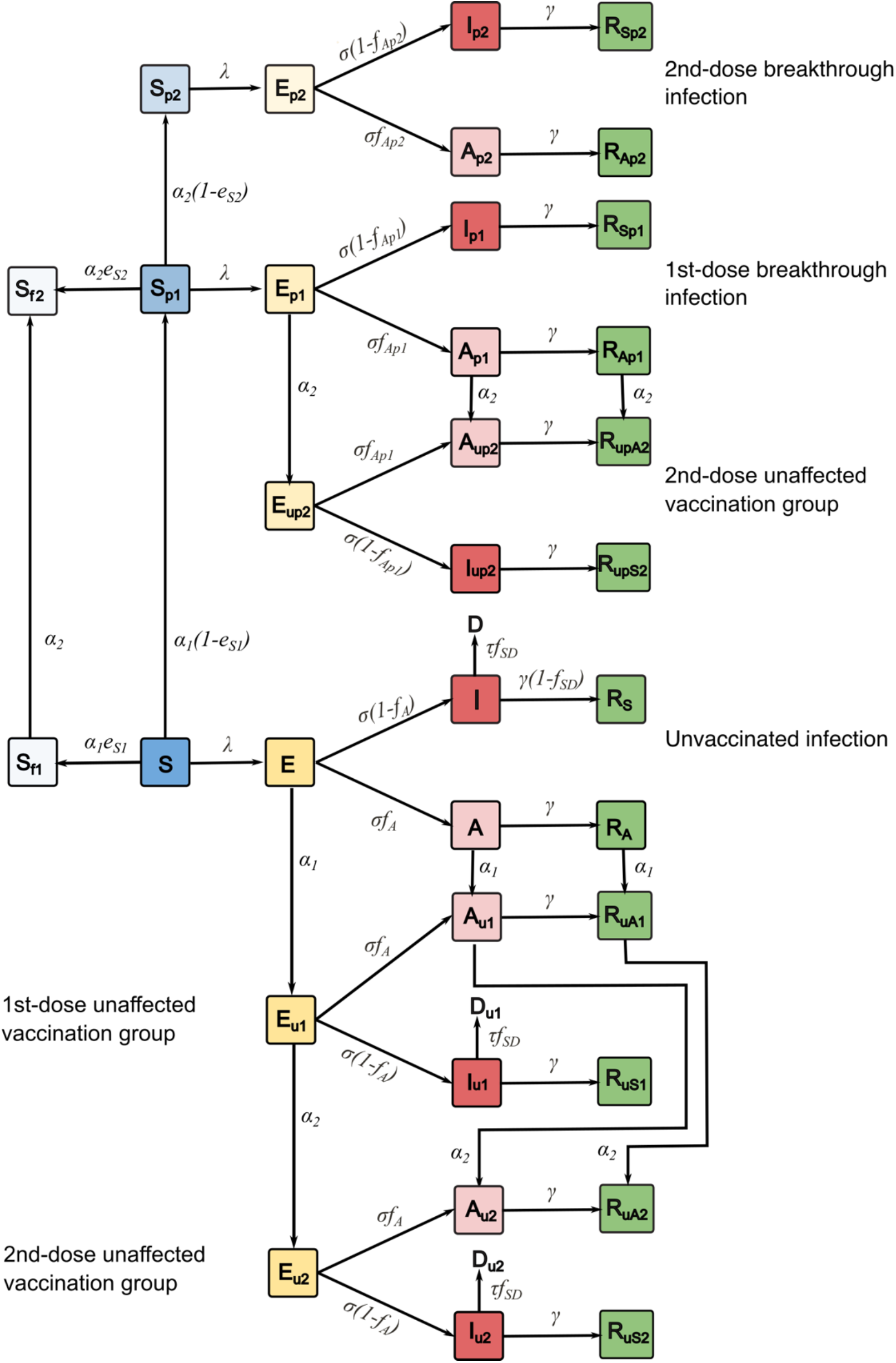
Schematic of COVID-19 transmission model with two-dose vaccination.

We assumed that only individuals in the S, E, A, and RA compartments could be vaccinated; however, vaccine-induced immunity only plays a role in vaccinated susceptible individuals. Vaccinated individuals in the E, A, and RA compartments will be protected by infection-induced immunity [21]. After susceptible individuals are vaccinated, they move to either a fully (Sf) or partially (Sp) infection-protected compartment with the ratio of Sf and Sp determined by the efficacy against infection of the vaccine (*eS*). When individuals in the partially infection-protected compartment get infections, they will move to the exposed compartment Ep. The parameters *α*_1_ and *α*_2_ are the rollout speeds per capita for the first and the second dose of vaccines, respectively.

The force of infection, *λ*_*j*_, for individuals in agee group *i* is given by

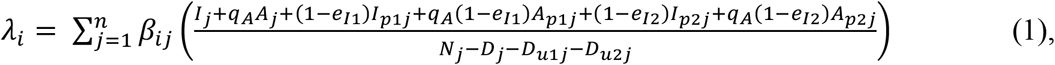

where *β*_*ij*_ is the transmission rate from infectious individuals in age group *i* to susceptible individuals in age group *j*, which is proportional to the contact frequency (*c*_*ij*_) [22]. *qA* is a parameter representing the relative infectiousness of asymptomatic individuals compared to that of symptomatic ones. The vaccine efficacies against infection, transmission, and symptomatic disease from the first dose of vaccination are denoted as *e*_*S*1_, *e*_*I*1_ and *e*_*D*1_ respectively. *e*_*S*2_, *e*_*I*2_ and *e*_*D*2_ are the additional vaccine efficacies induced from the second vaccination dose. The terms (*1 – eI1*) and (*1 – eI2*) represent the transmission contributed from the infectious individuals who have already been vaccinated for one and two doses, respectively. *Nj* is the total number of individuals in age group *j*, and *Dj* is the number of individuals in age group *j* who have died. The simulations were performed and analyzed using MATLAB R2020b.

The vaccination and transmission dynamics are described by the following systems of differential equations, where *i* = 1, …, 16 refers to the age groups of the populations:

Unvaccinated infections:

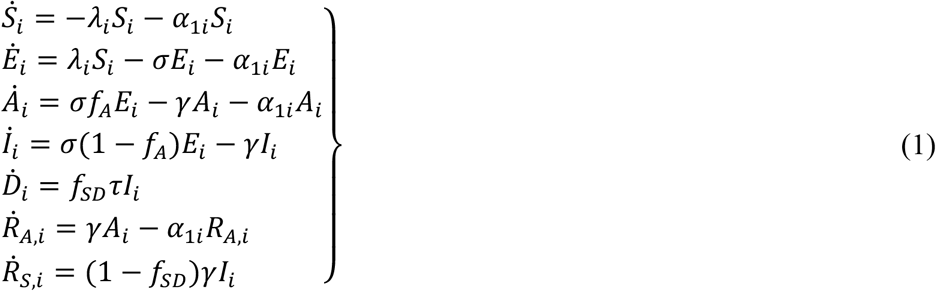

1^st^-dose breakthrough infections:

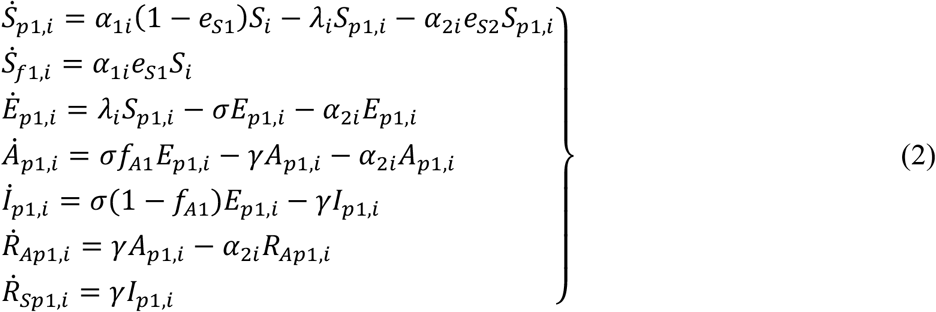

2^nd^-dose breakthrough infections:

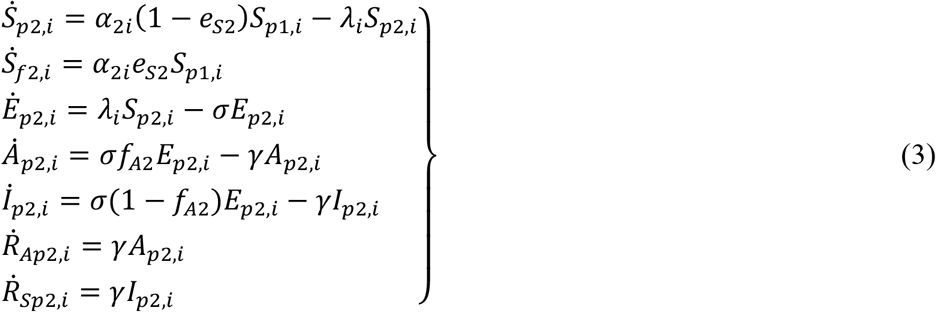

Unaffected vaccination groups:

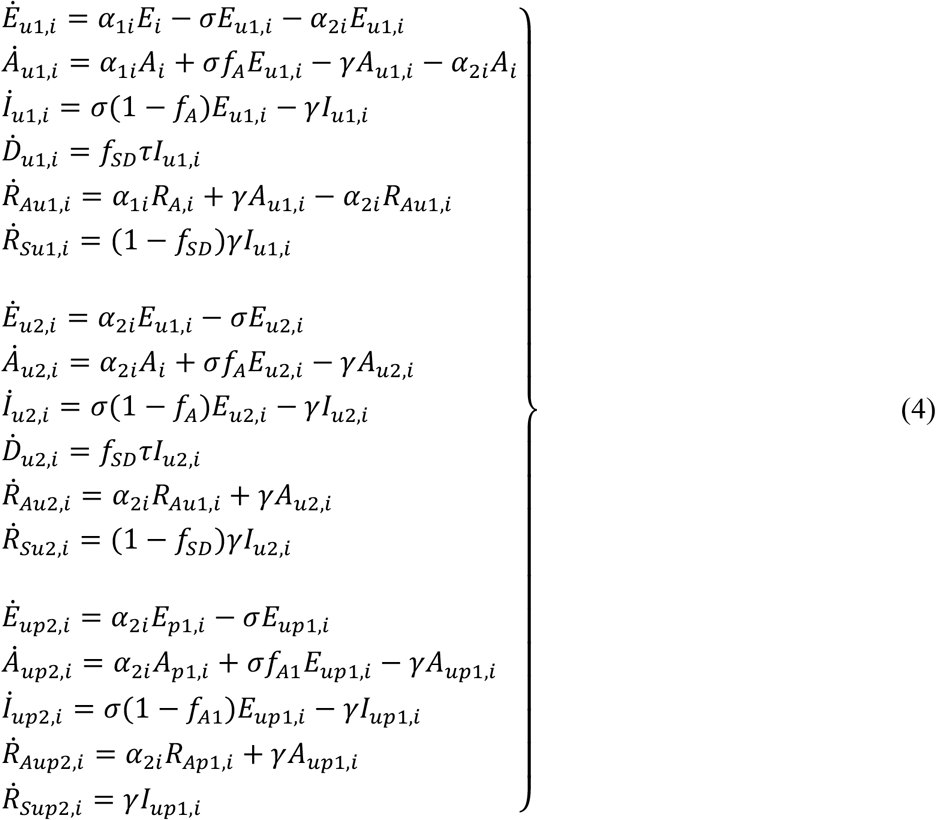

All parameters and their values used in the model are summarized in **Table 1**.

**Table 1.**
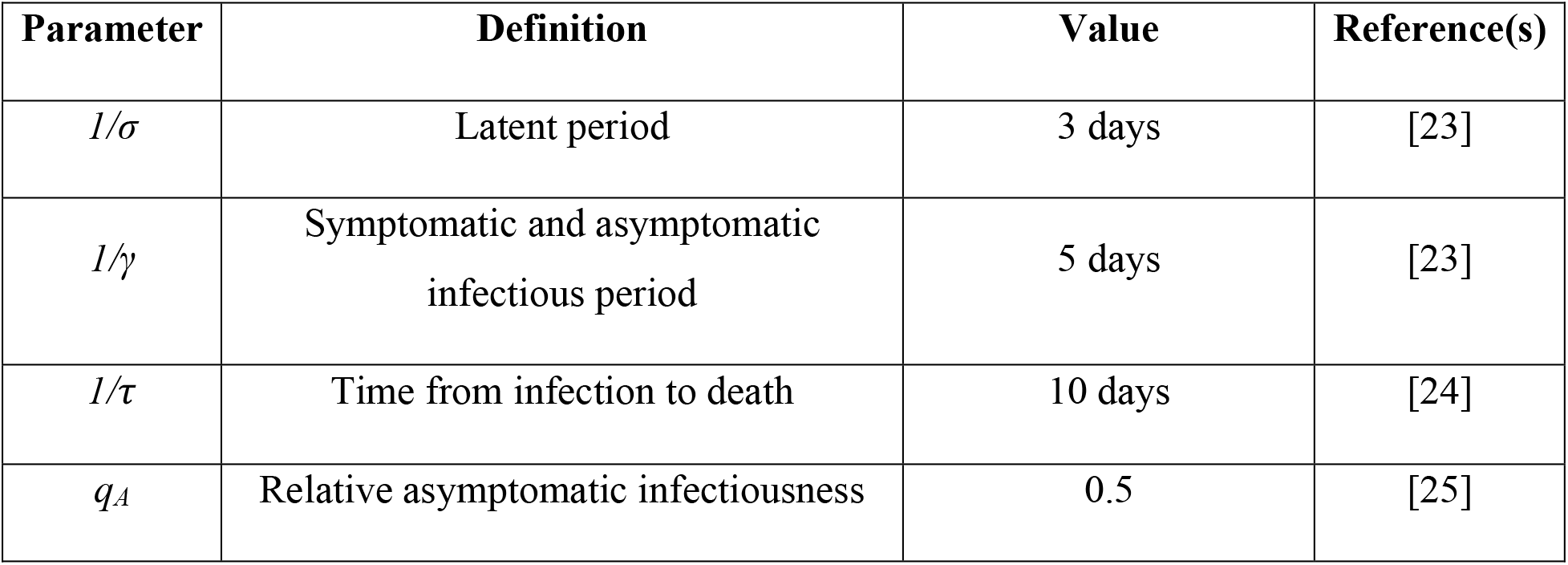
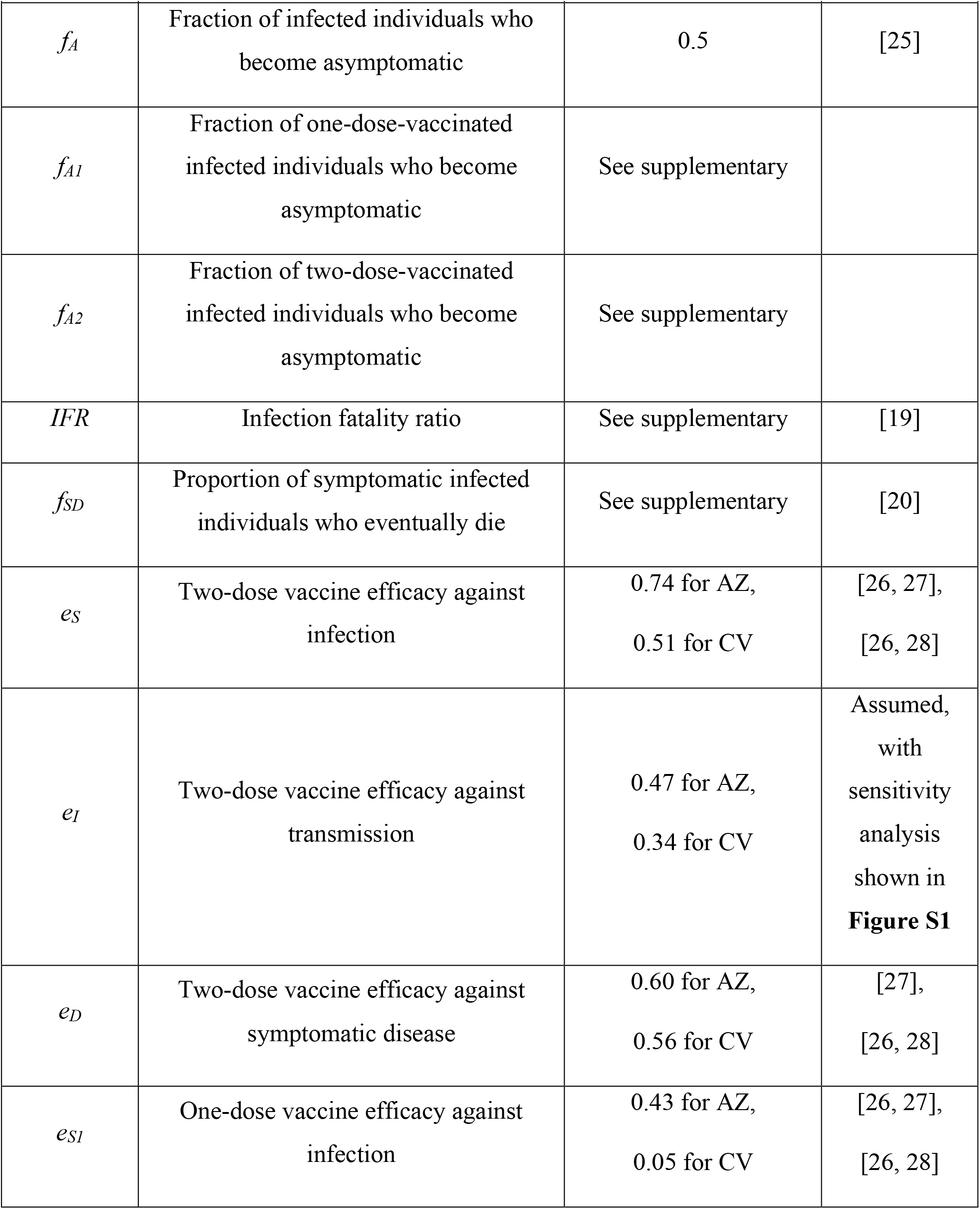
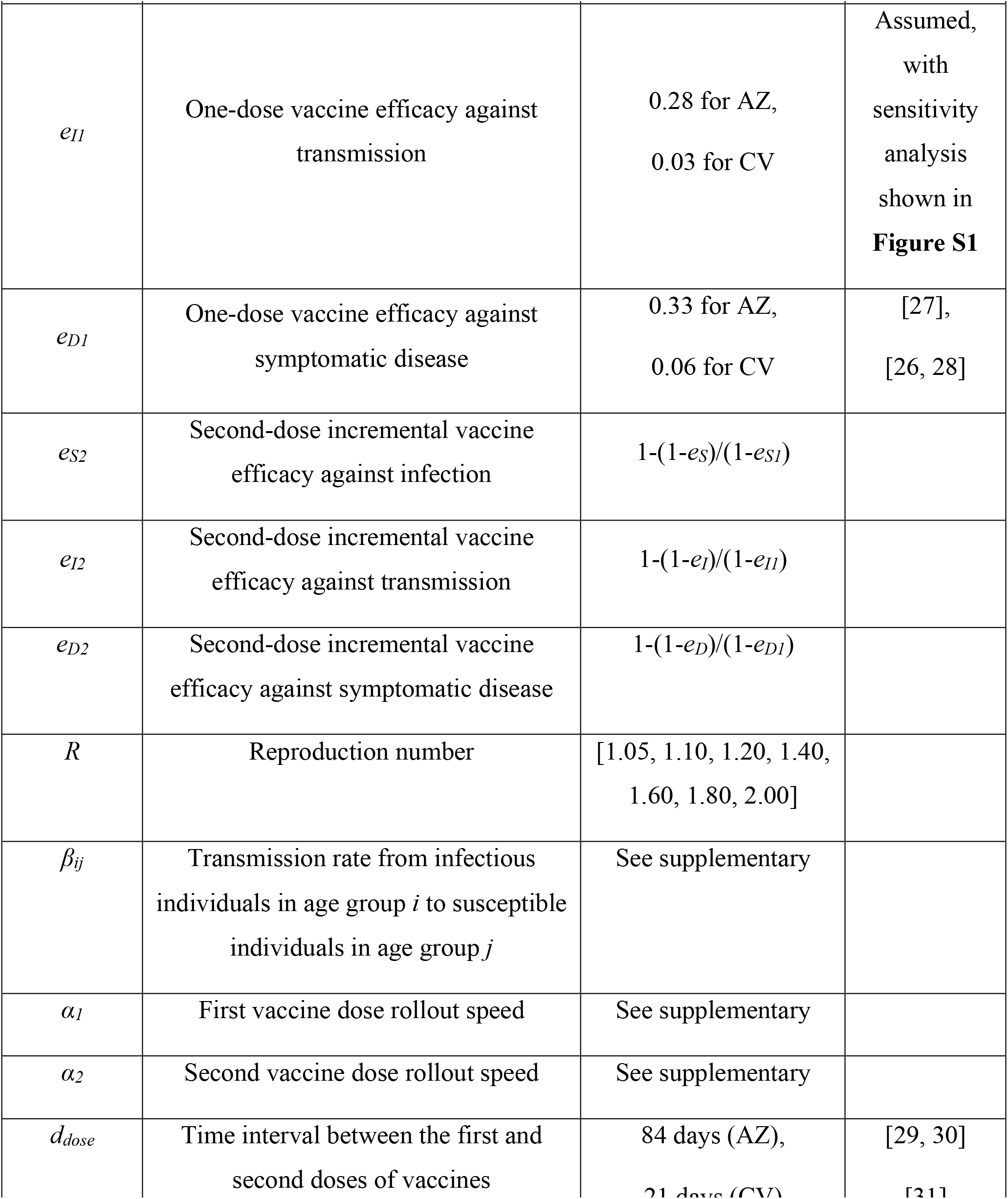
Parameters and their default values used in the model.

### Homologous and heterologous vaccination strategies

We considered three vaccination strategies for homologous vaccination, namely, homologous CV vaccination (CV+CV), homologous AZ vaccination (AZ+AZ), and parallel homologous CV and homologous AZ vaccination (CV and AZ parallel). The vaccines can be distributed to two target groups, namely, workers (20-60 years) and elders (≥ 60 years), with different prioritization strategies. The vaccination starts from the beginning of the simulation (*t* = 0). According to the vaccination roadmap announced by the Thai government in July 2021 [32], the vaccine rollout speed was set at a constant speed of 10 million doses/month. The recommended time intervals between the first and second doses of the vaccines (*d*_*dose*_) were set to 21 and 84 days for the CV vaccine [31] and the AZ vaccine [29], respectively.

For the heterologous vaccination strategy, since the first available COVID-19 vaccine in Thailand was the CV vaccine, we, therefore, investigated the scenario where the CV vaccine is used as the first dose, followed by the AZ vaccine as the second dose with the dosing interval (*d*_*dose*_) of 21 days [33, 34]. As the individuals who got the CV-AZ heterologous vaccination have a similar level of immunity as the homologous AZ-AZ homologous vaccination [33], we assumed that the vaccine efficacies of the one-dose and two-dose heterologous vaccination correspond to the efficacies of the one-dose of CV and two-dose of AZ vaccine, respectively. In these heterologous vaccination scenarios, the disease transmission was assumed to start when the AZ vaccine was available.

To accelerate vaccination, all vaccine available initially was assumed to be delivered as the first dose without reserving for the second dose during the first day of vaccination (*t* = 0) to the last day of the time interval (*t* = *d*_*dose*_) [35]. After that, all vaccine supplies would be administered as the second dose. The vaccination would return to the first dose again when no more individuals were due for the second dose. The alternating vaccination of first and second doses was repeated until the desired vaccination coverage was achieved. The vaccine rollout speed for the first (*α*_1_) and second (*α*_2_) doses for each time step was calculated from the ratio of the rollout speed and the number of individuals to be vaccinated (see **Supplementary B**).

### Prioritization strategies

To investigate the vaccine prioritization strategies in Thailand, we considered three different prioritization strategies: (i) no prioritization, where all individuals are randomly vaccinated, (ii) elder prioritization, and (iii) worker prioritization. Once the prioritized population was all vaccinated, vaccines were distributed to other target groups. In this part of the study, the homologous AZ vaccination (AZ+AZ) with rollout speeds of 50,000, 100,000, 250,000, and 500,000 doses per day were considered.

## Results

### Vaccination and COVID-19 situations in Thailand

Since the first infected case of COVID-19 was detected in Thailand in early 2020, two waves of the epidemic were well managed and controlled, with 29,127 cumulative cases and 95 deaths as of March 29, 2021. Early April 2021, the third wave of transmission began with the imported Delta variant [12, 36]. The Delta variant led to rising confirmed reported cases from February 28 to November 31, 2021 (**Figure 2a**). The maximum number of daily confirmed cases was up to 21,838 cases per day on August 7, 2021. The peak of daily deaths in Thailand occurred on August 18, 2021, with 312 confirmed COVID-19 deaths (**Figure 2b**). The lockdown measure was implemented from July 20 to November 1 [34], as shown in the yellow highlight in **Figure 2a** and **2b**.

**Figure 2.**
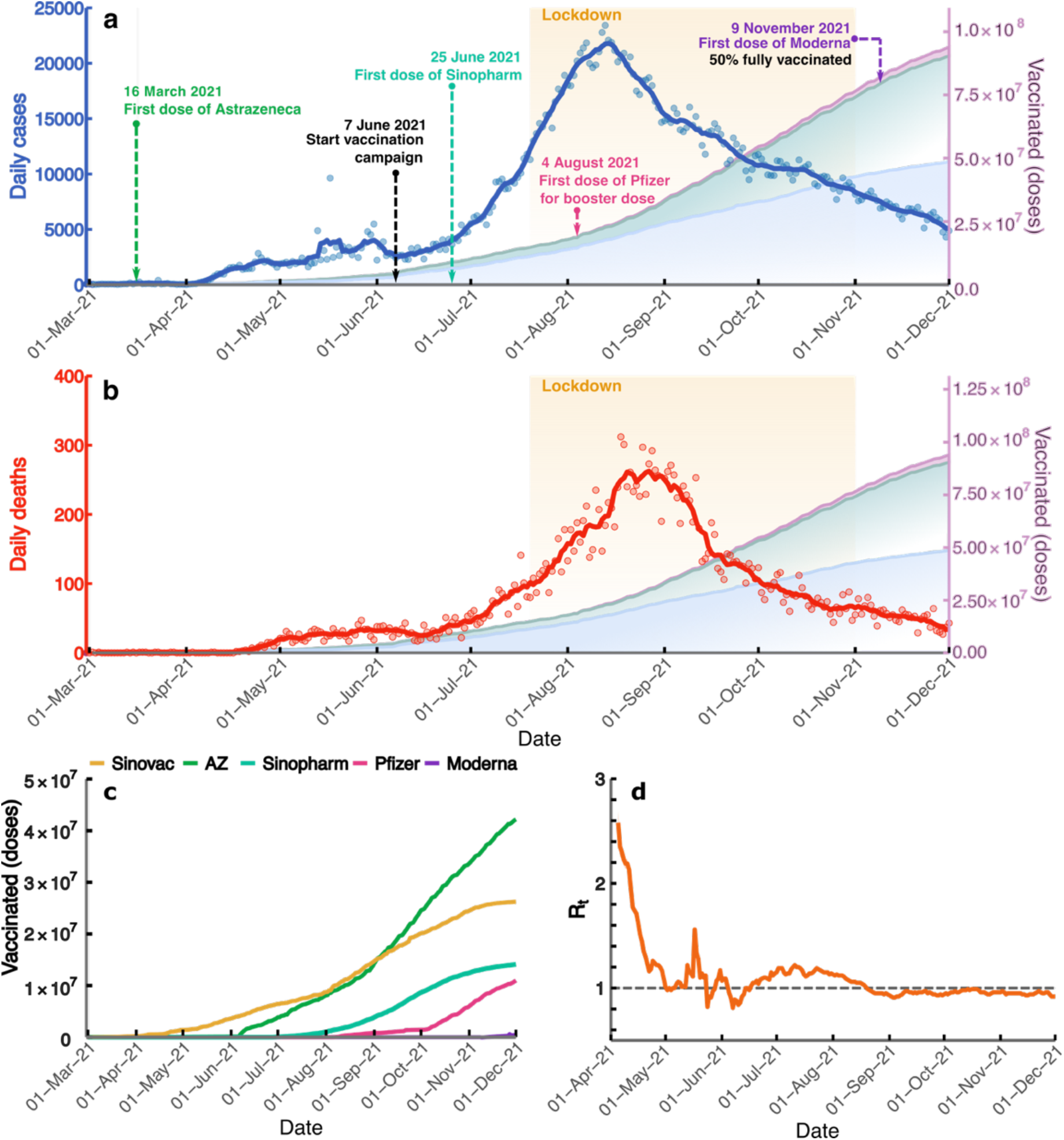
Vaccination and COVID-19 situations in Thailand. The blue line **(a)** and the red line **(b)** illustrate the number of daily new cases and the number of daily deaths, respectively. The blue, green, and purple areas show the cumulative numbers of first, second, and booster doses, respectively. The black-dashed arrows indicate the time point when the vaccination campaign started. The colored dashed arrows point to the start date of administering the vaccines from the different manufacturers. The yellow highlight area shows the duration of the lockdown measure. **(c)** Cumulative doses of vaccines from different manufacturers that have been delivered. **(d)** The time-varying reproduction numbers starting from April 1 to December 1, 2021. The line shows the median of *R*t, and the shaded area indicates the 95% CI. The horizontal dashed line indicates the *R*t threshold value of 1.

Regarding COVID-19 vaccination in Thailand, the first doses of CV and AZ vaccines were first delivered to the healthcare workers on February 28 and March 16, respectively, while the national vaccination campaign started on June 7. Later, on July 25, the Sinopharm BBIBP vaccines were imported by the Chulabhorn Royal Academy for sale and charity [37]. The Comirnaty from Pfizer-BioNTech COVID-19 vaccine, donated by the United States, was used as booster doses for healthcare workers who got the two doses of CV vaccine and for children aged 12-18 years. Additionally, Spikevax from Moderna, imported by a private hospital in Thailand, was first administered on November 9, when the transmission wave had already gone down. Due to the shortage of vaccines at the beginning of the vaccination campaign in Thailand, many strategies of the heterologous vaccination were employed; the CV vaccine was used as the first dose and AZ vaccine as the second dose. At the end of 2021, around 44 million, 26.4 million, and 17.1 million doses of AZ, CV, and Pfizer were administered, respectively. The numbers of cumulative first (blue), second (green), and booster (purple) doses are shown in **Figures 2a** and **2b**.

We estimated the time-varying reproduction number (*Rt*), which is the average number of secondary cases generated by an infected person. A value of *Rt* greater than the threshold value of 1 indicates that the number of new infections is growing at time *t*, whereas *Rt* less than 1 indicates that the epidemic size is shrinking at time *t*. We employed a method proposed by Cori and colleagues [36, 38] to estimate the *Rt* in Thailand. *Rt* was calculated starting from April 5, when the number of daily cases exceeded 100, to December 1, 2021 (**Figure 2d**). We found that the median *Rt* in the early phase of transmission was 2.57 (95% CI: 2.36-2.80). However, Thailand implemented lockdown measures in early April 2021, resulting in the *Rt* decreasing to a value below one during that time. In July 2021, the number of deaths and cases drastically increased due to the spread of the more transmissible Delta variant. As a result, the *Rt* value rose to a value greater than one. The Thai government then expanded the lockdown areas to other parts of the country and increased the vaccination speed [39]. After Thailand launched the national COVID-19 vaccination program, the *Rt* decreased to a value below one on August 26, 2021, and has since remained below one until December 1, 2021.

### Impacts of homologous and heterologous vaccination strategies on mortality and infections

We investigated the effect of homologous and heterologous vaccination strategies on the reduction in cumulative numbers of deaths and cases. In this study, there were three scenarios for the homologous vaccination strategy: i) homologous CV (CV+CV), ii) homologous AZ (AZ+AZ), and iii) parallel homologous CV and homologous AZ vaccinations (CV and AZ parallel). For the heterologous vaccination, we investigated the strategy in which the population gets the CV vaccine as the first dose and the AZ vaccine as the second dose. **Figure 3** shows the reduction in the cumulative number of deaths due to different vaccination strategies. For the homologous vaccination strategies, we found that the AZ+AZ strategy can advert more deaths than the CV+CV strategy across the entire range of reproduction numbers (**Figures 3f)**. However, in the scenarios where the CV and AZ vaccines are both employed, we found that the CV+AZ heterologous vaccination performs better than the parallel CV and AZ homologous vaccination, especially when the effective reproduction number is lower than 1.4 (**Figures 3f)**. Similar results were also found in terms of reducing the number of cumulative cases (**Figure 4**). Note, however, that since both CV and AZ vaccines were concurrently employed in CV+AZ heterologous and parallel CV and AZ homologous vaccination strategies, the vaccination rate for these two strategies is, therefore, two times higher than that of the homologous CV and homologous AZ vaccination strategies.

**Figure 3.**
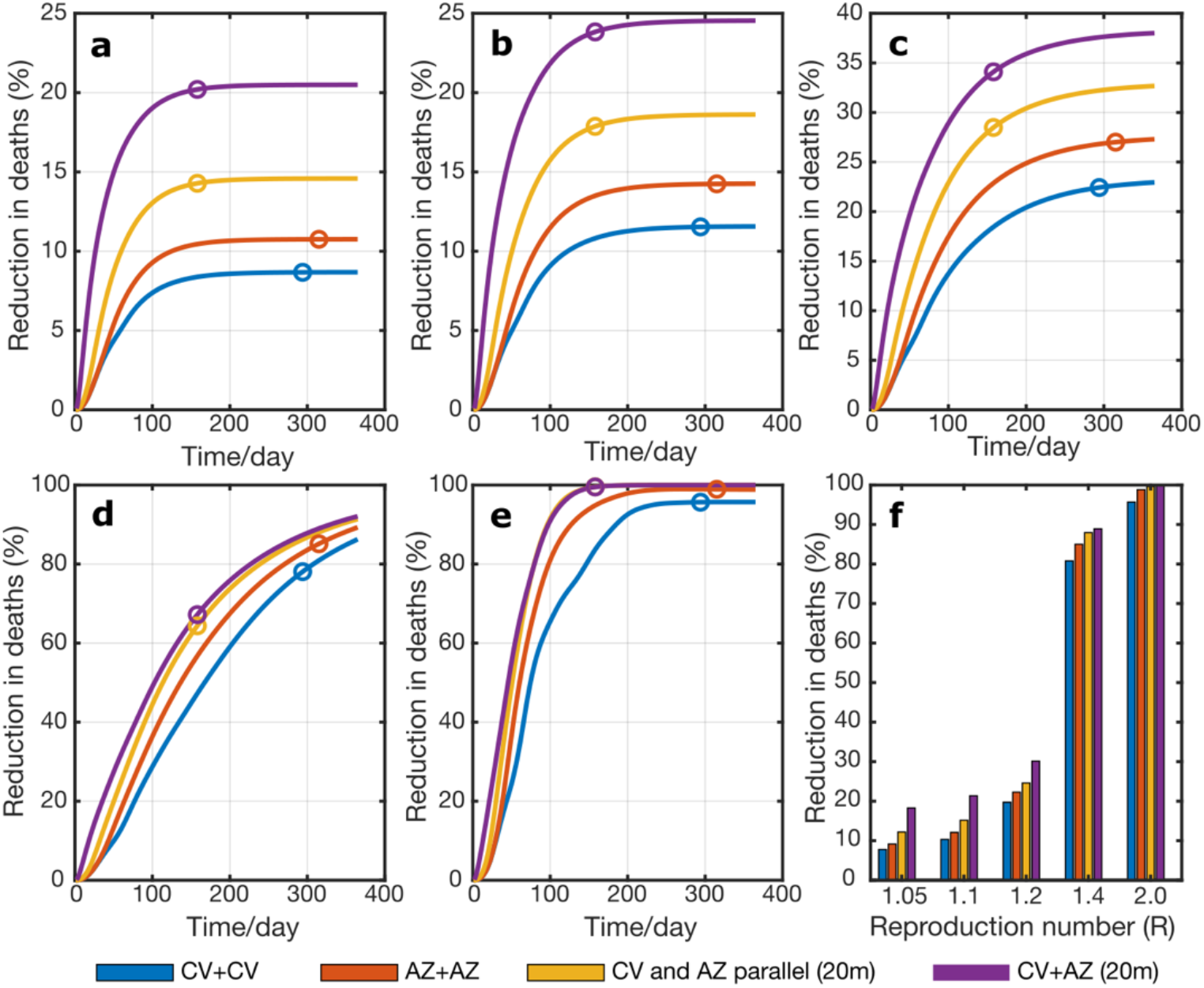
Reduction in cumulative deaths under different vaccination strategies. Results were obtained from model simulations with the reproduction number of **(a)** *R* = 1.05, **(b)** 1.1, **(c)** 1.2, **(d)** 1.4, and **(e)** 2.0 with either CV+CV (blue), AZ+AZ homologous vaccinations (red), parallel homologous vaccination of CV and AZ (yellow), and CV+AZ heterologous vaccination (purple). Color dots represent the times at which 70% of individuals were vaccinated. **(f)** The comparison of the reduction in deaths at the equilibrium under different vaccination scenarios.

**Figure 4.**
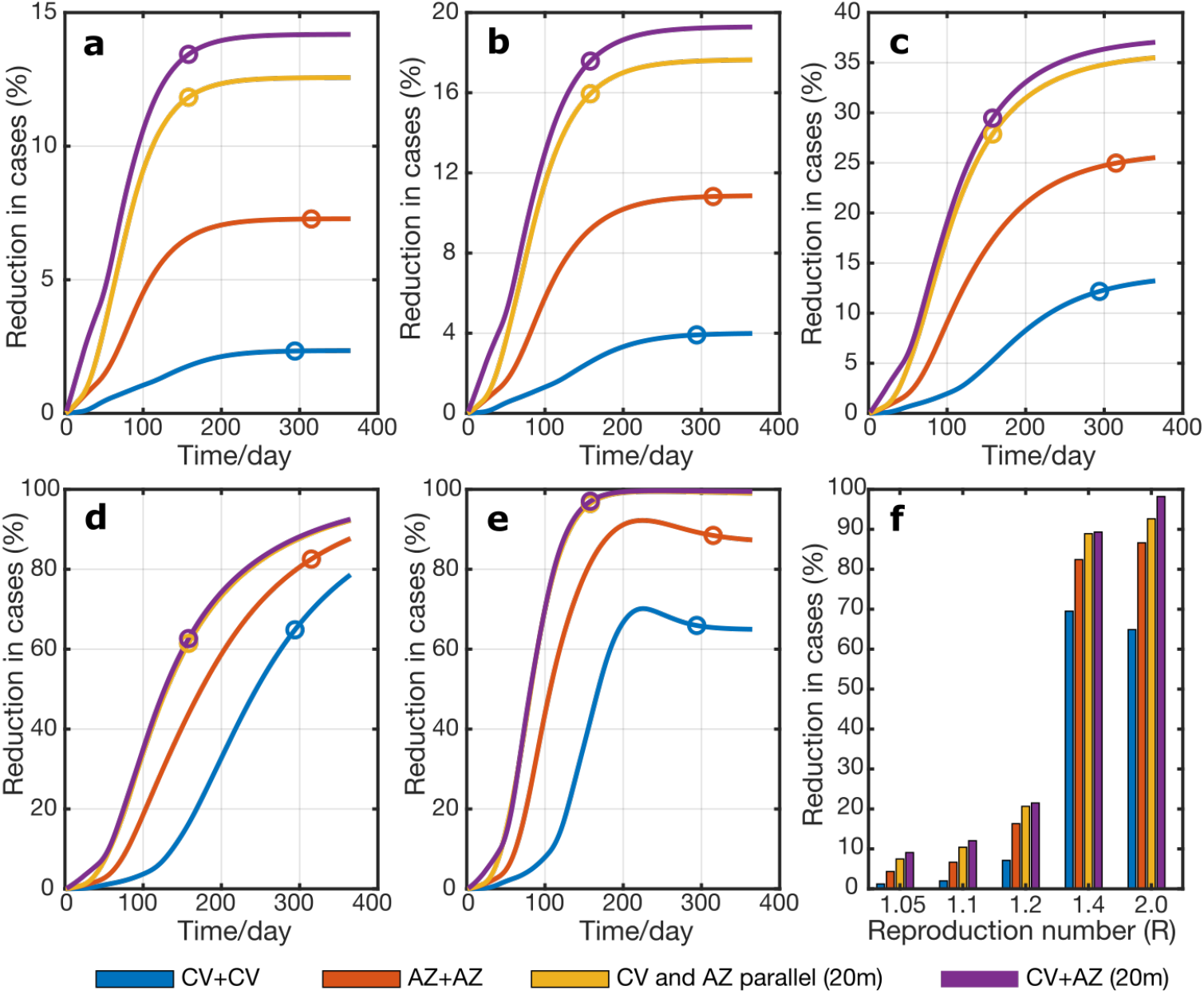
Reduction in cumulative cases under different vaccination strategies. The cumulative numbers of cases were from model simulations with the reproduction number of **(a)** *R* = 1.05, **(b)** 1.1, **(c)** 1.2, **(d)** 1.4, and **(e)** 2.0 with either CV+CV (blue), AZ+AZ homologous vaccinations (red), parallel homologous vaccination of CV and AZ (yellow), and CV+AZ heterologous vaccination (purple). Color dots represent the times at which 70% of individuals were vaccinated. **(f)** The comparison of the reduction in cumulative cases at the equilibrium under different vaccination scenarios.

### Impact of vaccine prioritization strategies and vaccine rollout speeds

We investigated three different vaccine prioritization strategies: no priority (vaccinate individuals of all ages simultaneously), elderly priority (aged 60 years and older), and worker priority (aged 20-59 years). Vaccines were first distributed to all individuals in the priority age group. After all individuals in the priority age group were vaccinated, the vaccines were allocated to other age groups. The impact of vaccine prioritization strategies was determined by a reduction in cumulative numbers of deaths and cases at the equilibrium (*t* = 2000 days). Vaccines were distributed at a rate of 50,000, 100,000, 250,000, and 500,000 doses/day, corresponding with a range of vaccination capacity in Thailand. The homologous AZ+AZ vaccination was employed in this part of the study.

Figure 5 shows the impacts of different prioritization strategies on mortality and infections. We found that a faster vaccine rollout speed could result in a higher reduction in mortality and infection for all prioritization strategies. For a high-supply scenario with a rollout speed of 500,000 doses/day, the elderly priority strategy was found to be more effective in reducing deaths compared to other strategies. In contrast, for a low-supply scenario with a rollout speed of 50,000 doses/day, prioritization of vaccines to the elderly age group resulted in the smallest effect on reducing infections and deaths (**Figure 5b** and **5e**). In this scenario, the reduction in cumulative cases is maximized when vaccines are firstly allocated to the working age group.

**Figure 5.**
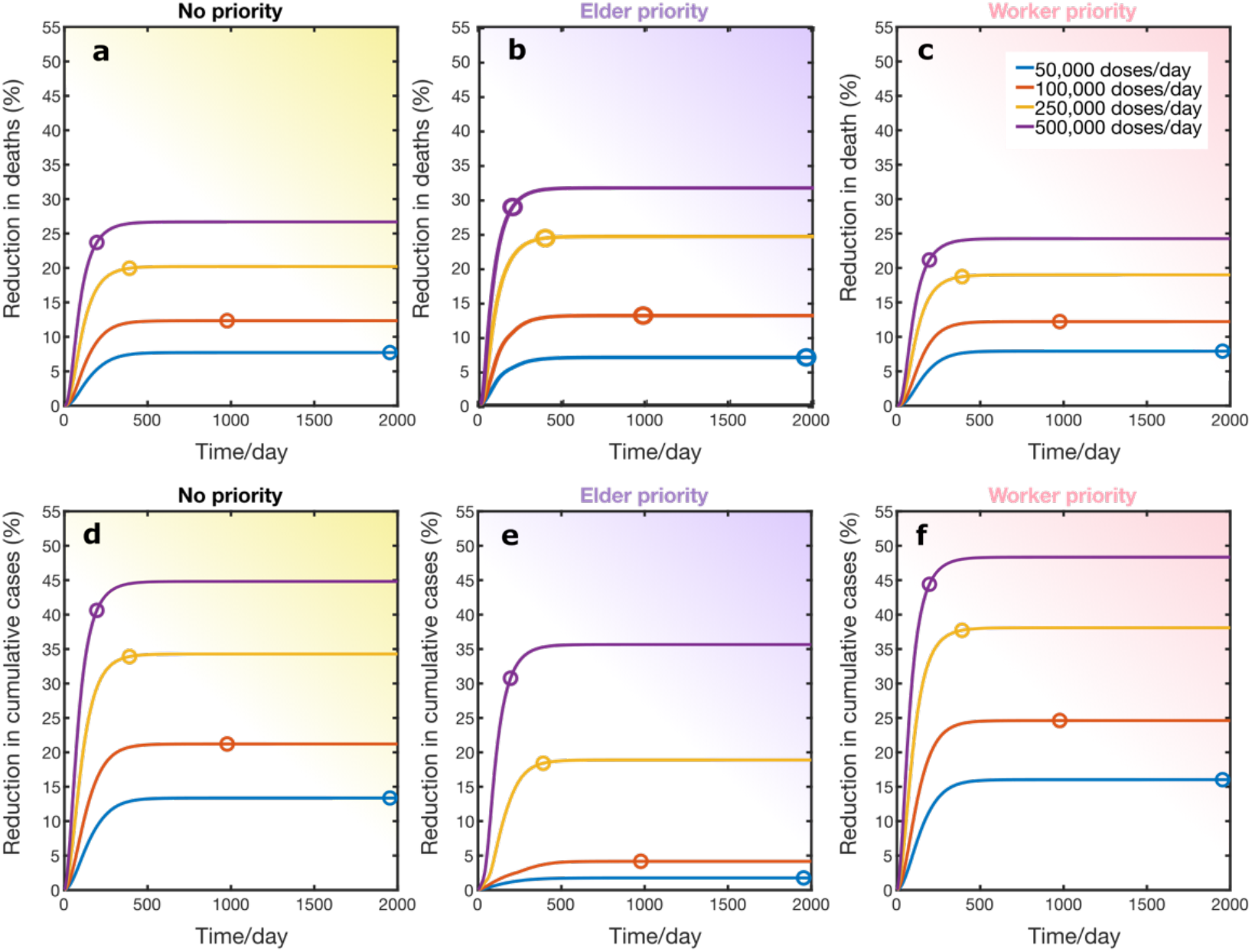
Impact of vaccine prioritization strategies on mortality and infections. **(a-c)** Reductions in cumulative deaths and **(d**-**f)** reductions in cumulative cases simulated using *R* = 1.2 under different vaccine rollout speeds. (**a, d**), (**b, e**), and (**c, f**) show the results under the random vaccine distribution (no priority), elderly priority, and worker priority, respectively. Color dots show the times when 70% of the total population was fully vaccinated.

To investigate the impact of the vaccine prioritization strategies on mortality and infections under different disease transmission dynamics, we varied the reproduction numbers and measured the percentage reductions in cumulative deaths and cases (**Figure 6**). We found that for slow transmission dynamics (*R* = 1.05, 1.1, and 1.2), prioritizing the elderly aged 60 years and above can reduce COVID-19 mortality more than other strategies for all vaccine rollout speeds (**Figure 6a**-**c**). For the disease transmission with *R* = 1.4, all vaccine prioritization strategies reduced mortality similarly (**Figure 6d**). In this case, the reductions in deaths for the elderly and worker prioritization strategies were found to be 97% and 98%, respectively. For very fast transmission dynamics (*R* = 2.0), we found that vaccinating the elderly first could advert more deaths when the vaccine rollout speed is low (< 100,000 doses/day) (**Figure 6e**). However, in a high-supply scenario with a rollout speed of 250,000-500,000 doses/day, all vaccine prioritization strategies were similar in reducing mortality. In contrast, the results revealed that vaccinating individuals in the working age group firstly always most reduced the number of cases (**Figure 6f** to **6j**).

**Figure 6.**
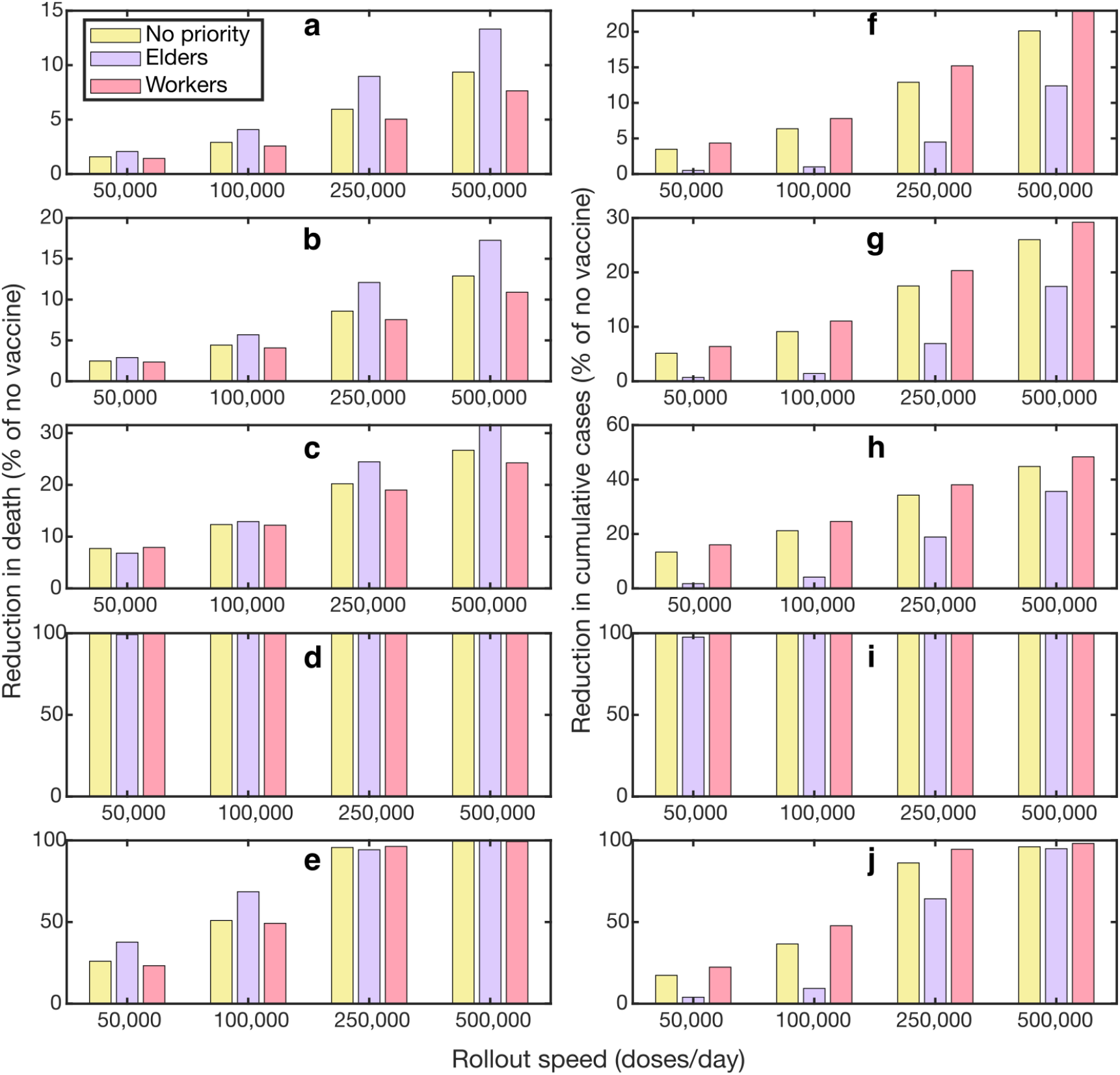
Effects of vaccine prioritization strategies on COVID-19 mortality and infections. **(a-e)** Percentage reduction in cumulative deaths for varying vaccine rollout speeds in the scenario where *R* = 1.05, 1.1, 1.2, 1.4, and 2.0, respectively. **(f-j)** Percentage reduction in cumulative cases for varying vaccine rollout speeds in the scenario where *R* = 1.05, 1.1, 1.2, 1.4, and 2.0, respectively.

## Discussion

During the COVID-19 pandemic, early access to COVID-19 vaccines in low-income countries was very limited. At the end of 2021, less than 10% of the low-income countries’ population received at least one dose of the vaccine, while most people in wealthier countries were fully vaccinated, constituting the vaccine inequity problem [12, 40-42] (**Figure S2**). The COVID-19 vaccination in Thailand started on February 28, 2021. In this research, we delineated the timeline of COVID-19 vaccination during the first nine months of COVID-19 vaccination in Thailand (**Figure 2**). During the early phase of vaccination, the CoronaVac (CV) was the only vaccine available in Thailand partly due to the delay in delivery of ChAdOx1 nCoV-19 by AstraZeneca plc (AZ). Most of the early available vaccine doses were allocated to healthcare workers, the elders, and high-risk groups. The national vaccination campaign in Thailand started on June 7, 2021 [43], when the Delta variant began to circulate in Thailand, and the number of cases and deaths continued to increase. To control SARS-CoV-2 transmission under the limited vaccine supply, the heterologous vaccination strategy, in which the CV vaccine was used as a first vaccination dose and AZ as a second dose, was then introduced [44].

To assess the effectiveness of the heterologous vaccination strategy employed in Thailand, the age-structured model of COVID-19 transmission and vaccination was constructed. The model allowed us to directly compare to heterologous vaccination strategy to the traditional homologous vaccination under different disease transmission dynamics. The reproduction number was assumed to have a value in the range of 1.05 and 2.00, reflecting the COVID-19 transmission under control measures in Thailand (**Figure 2d**). By measuring the cumulative numbers of cases and deaths averted by vaccination, we found that the CV+AZ heterologous vaccination strategy performs better than the separate CV and AZ homologous vaccinations in reducing the cumulative numbers of cases and deaths, especially when other nonpharmaceutical interventions can suppress the effective reproduction number to lower than 1.4 (**Figures 3f)**. This might be due to the fact that the time interval between the first and the second dose for the CV+AZ heterologous vaccination is shorter than that of the AZ homologous vaccination.

The duration between two doses of the AZ homologous vaccination is recommended to be four to twelve weeks apart as the vaccine efficacy tends to be higher when the duration between doses is longer [45, 46]. Hence, most people in Thailand got the second AZ dose twelve weeks after their first dose, which could slow down the number of fully vaccinated individuals in the community. However, if individuals get the CV vaccine as their first dose, they can get the second dose three weeks after the first dose. Hence, the dosing interval for the CV+AZ heterologous vaccination can be shortened to three weeks, compared to twelve weeks in AZ homologous vaccination, while maintaining the same vaccine efficacy [47, 48]. This CV+AZ heterologous vaccination strategy could, therefore, accelerate the number of fully vaccinated individuals. The results also indicated that the faster the heterologous vaccine rollout speed, the greater the reduction in both deaths and cases. These results suggested that for low- and middle-income countries where early access to high-efficacy vaccines is limited, getting any affordable vaccines as early as possible and using them as the first dose for heterologous vaccination might be a better strategy for preventing a severe outcome [49].

We also investigated the impact of the vaccine prioritization strategies on COVID-19 mortality and infections. Our results suggested that a faster vaccine rollout speed could rapidly reduce more COVID-19 infections and deaths in all vaccine prioritization strategies. Thus, the fast rollout speed of the vaccine is critical to quickly curbing the spread of COVID-19. Moreover, we found that the most effective vaccine prioritization strategy depends on the reproduction number of the COVID-19 transmission. For the slow transmission dynamics (maybe due to intense non-pharmaceutical interventions), prioritizing the elderly can reduce COVID-19 mortality more than other strategies for all vaccine rollout speeds. In the scenario where the transmission dynamics are fast (*R* = 2.0) and the rollout speed of the vaccine is fast (in the range of 250,000-500,000 doses/day), all vaccine prioritization strategies were equally effective at reducing mortality. However, if the vaccine rollout speed is slow (in the range of 50,000-100,000 doses/day), vaccinating the elderly first can drastically reduce mortality more than other strategies (**Figure 6**). Indeed, Thailand has implemented a very strict lockdown combined with case isolation and non-pharmaceutical interventions such as physical distancing and face masking during the COVID-19 outbreak [12, 36, 50]. This has resulted in a low effective reproduction number during the outbreak (**Figure 2d**). As a result, vaccination strategies targeting the elderly in Thailand could be an optimal strategy in terms of reducing COVID-19 mortality.

In terms of vaccine prioritization to reduce COVID-19 infections, our results suggested that prioritizing vaccines among workers (aged 20-59 years) could lower the number of COVID-19 cases more than the other strategies. Our results are broadly consistent with other studies that recommended prioritizing essential workers to minimize COVID-19 cases [51-53]. This is because the workers have high contact frequencies [52, 54]. We found that the worker priority vaccination could reduce the number of COVID-19 cases at all values of the considered reproduction number *R* (**Figure 6f** to **6j**).

## Conclusion

In conclusion, we discovered that the CV+AZ heterologous vaccination strategy performs better than the separate parallel CV and AZ homologous vaccination strategy in adverting COVID-19 infections and mortality when paired with other intensive nonpharmaceutical measures. This is attributable in part to the shorter period between the first and second doses of the CV+AZ heterologous immunization compared to the AZ homologous vaccination. To minimize the impacts of disparities in early COVID-19 vaccine access in low- and middle-income countries, our modeling results suggested that these nations may employ early accessible but possibly lower-efficacy vaccines in combination with higher-efficacy vaccines for the first and second doses of heterologous vaccination, respectively. In addition, the findings revealed that prioritizing vaccines for the elderly may be best for reducing COVID-19 mortality over a broad range of vaccination rates and transmission dynamics.

## Data Availability

All data produced in the present work are contained in the manuscript.

## Acknowledgments

This study was financially supported by the Centre of Excellence in Mathematics, Ministry of Higher Education, Science, Research and Innovation, Thailand, Center of Excellence on Medical Biotechnology (CEMB), and Thailand Center of Excellence in Physics (ThEP).

## Supplementary Materials

### A. Vaccine rollout rate per population in each age group

We allowed vaccines to be rolled out at different rates by assigning the number of vaccine doses available each day, *rollout*. For each simulated day, vaccines were distributed, following the prioritization strategies. Firstly, only the first dose of the vaccine was delivered until the lead time, *d*_*dose*._ Then, the second dose of vaccine was delivered for the next lead time. After that, the vaccine distribution was repeated.

For the 1^st^ dose period (*t = [xd*_*dose*._*+1, (x+1) d*_*dose*._], x = 0, 2, 4, 6, …),

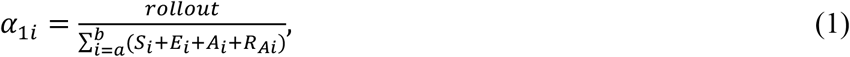

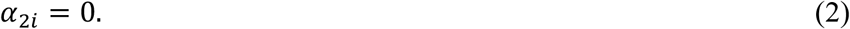

For the 2^nd^ dose period (*t =* [*xd*_*dose*._*+1, (x+1) d*_*dose*._], x = 1, 3, 5, 7, …),

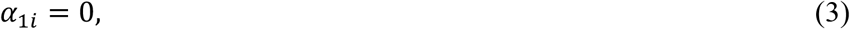

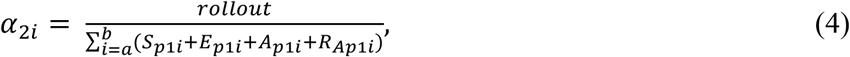

where *i* represents an age group. Note that the vaccines are not applied for the population below 20 years (group 1-4). *d*_*dose*._ for the CV vaccine and AZ vaccine were 21 and 84 days, respectively. *d*_*dose*._for the heterologous vaccine is 21 days. *a* and *b* are the age groups corresponding with the prioritization strategies: (i) no priority (*a* = 5, *b* = 16), elder priority (*a* = 13, *b* = 16), and worker priority (*a* = 5, *b* = 12).

### B. Estimate the fraction of asymptomatic breakthrough infections and the fraction of deaths

Since the efficacy against disease is the reduction in disease in a vaccinated group for the 1^st^ dose vaccine compared to an unvaccinated group,

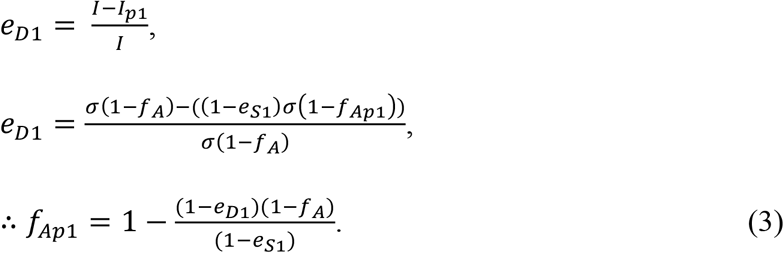

For the 2^nd^ dose vaccine, (*eD2* is calculated compared to an unvaccinated group)

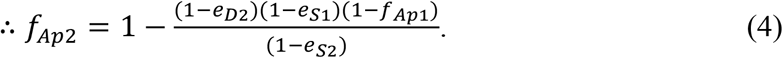

The fraction of deaths depends on the infection fatality ratio which is related to the age of the infected individuals as follows [1]:

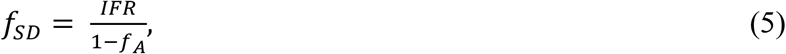

where *IFR* is the infection fatality ratio that relates to the age as

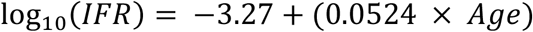

### C. Sensitivity analysis of the vaccine efficacy against transmission

**Figure S1.**
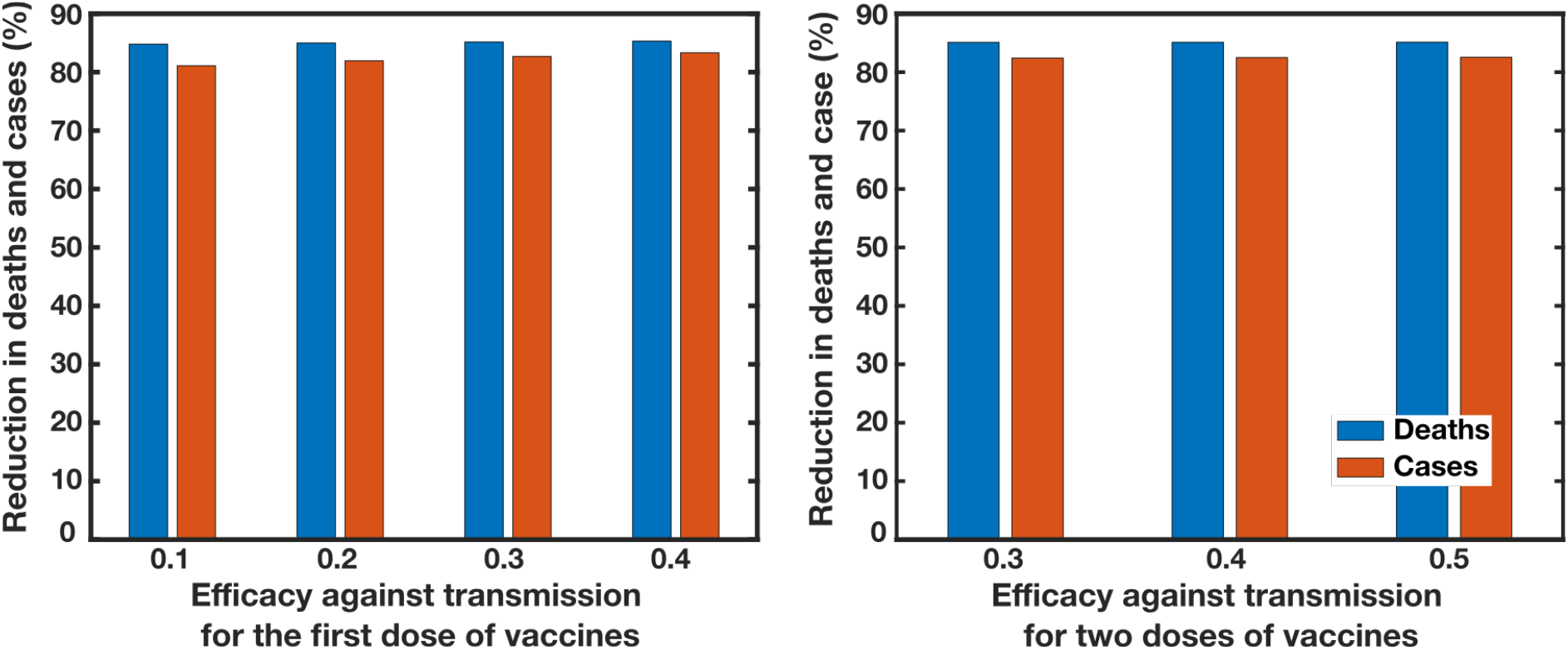
The reduction in cases (blue) and deaths (red) with (a) the variation of vaccine efficacy against transmission for the first dose, *eI1* where the vaccine efficacy of full vaccination = 0.47, and (b) the variation of *eI*, when *eI1* = 0.28.

### D. COVID-19 vaccine doses administered per 100 people

**Figure S2.**
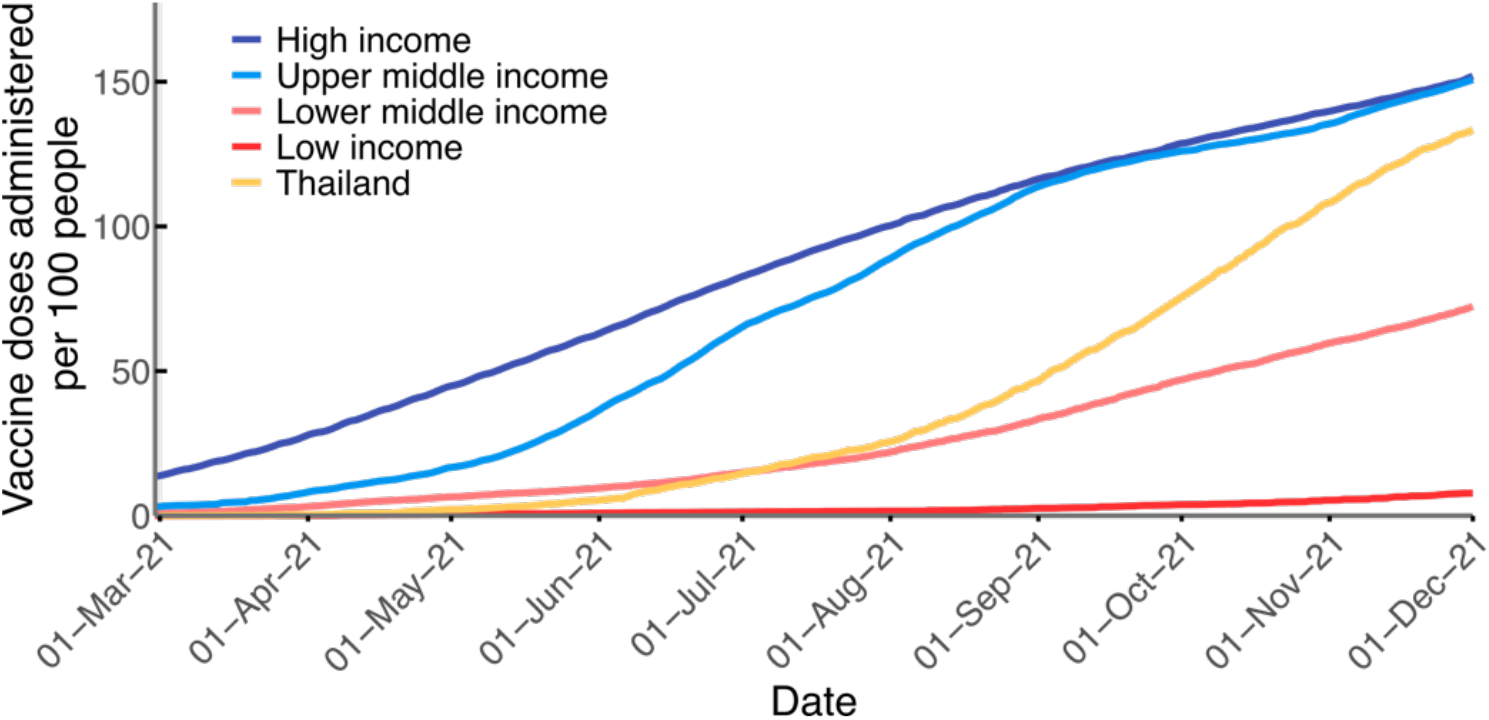
COVID-19 vaccine doses administered per 100 people by income groups. All doses, including boosters, are counted individually. As the same person may receive more than one dose, the number of doses can be higher than the number of people in the population [2].

### E. Effect of the prioritization strategies on the cumulative cases and deaths

**Figure S3.**
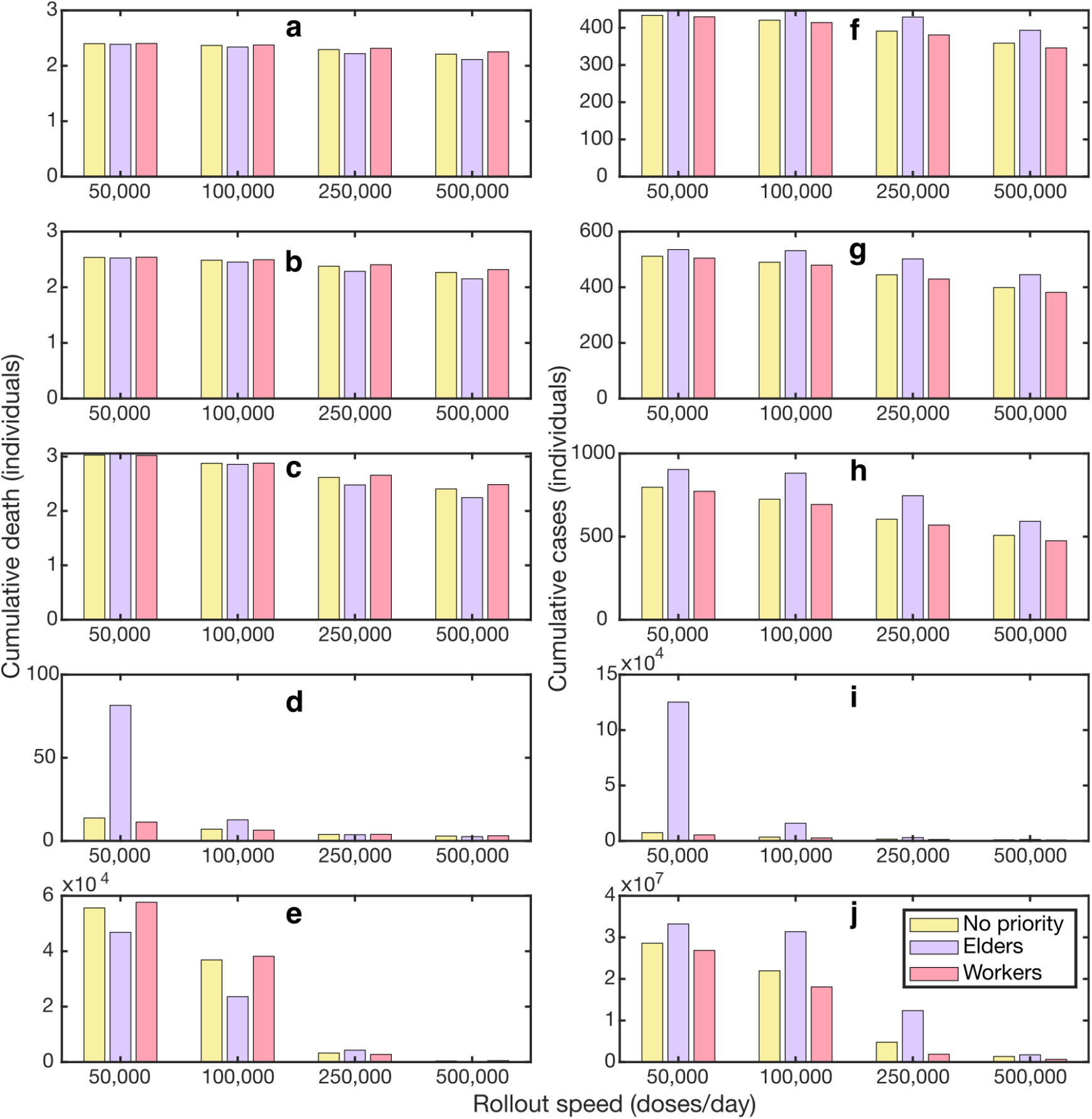
Effects of vaccine prioritization strategies on cumulative deaths and cases. **(a-e)** Cumulative deaths for varying vaccine rollout speeds in the scenario where *R* = 1.05, 1.1, 1.2, 1.4, and 2.0, respectively. **(f-j)** Cumulative cases for varying vaccine rollout speeds in the scenario where *R* = 1.05, 1.1, 1.2, 1.4, and 2.0, respectively.

## References

1. Zhu, N., et al., A Novel Coronavirus from Patients with Pneumonia in China, 2019. N Engl J Med, 2020. 382(8): p. 727–733.

2. Dong, E., H. Du, and L. Gardner, An interactive web-based dashboard to track COVID-19 in real time. Lancet Infect Dis, 2020. 20(5): p. 533–534.

3. Liu, Q., et al., Effectiveness and safety of SARS-CoV-2 vaccine in real-world studies: a systematic review and meta-analysis. Infectious Diseases of Poverty, 2021. 10(1): p. 132.

4. Lopez Bernal, J., et al., Effectiveness of Covid-19 vaccines against the B. 1.617. 2 (Delta) variant. N Engl J Med, 2021: p. 585–594.

5. Gardner, B.J. and A.M. Kilpatrick, Estimates of reduced vaccine effectiveness against hospitalization, infection, transmission and symptomatic disease of a new SARS-CoV-2 variant, Omicron (B.1.1.529), using neutralizing antibody titers. medRxiv, 2021: p. 2021.12.10.21267594.

6. Sheikh, A., et al., SARS-CoV-2 Delta VOC in Scotland: demographics, risk of hospital admission, and vaccine effectiveness. The Lancet, 2021. 397(10293): p. 2461–2462.

7. Higdon, M.M., et al., A systematic review of COVID-19 vaccine efficacy and effectiveness against SARS-CoV-2 infection and disease. medRxiv, 2021: p. 2021.09.17.21263549.

8. Andrews, N., et al., Covid-19 Vaccine Effectiveness against the Omicron (B.1.1.529) Variant. New England Journal of Medicine, 2022.

9. Hannah Ritchie, E.M., Lucas Rodés-Guirao, Cameron Appel, Charlie Giattino, Esteban Ortiz-Ospina, Joe Hasell, Bobbie Macdonald, Diana Beltekian and Max Roser, “Coronavirus Pandemic (COVID-19)". 2020: Published online at OurWorldInData.org.

10. Tregoning, J.S., et al., Progress of the COVID-19 vaccine effort: viruses, vaccines and variants versus efficacy, effectiveness and escape. Nature Reviews Immunology, 2021. 21(10): p. 626–636.

11. Maxmen, A. The fight to manufacture COVID vaccines in lower-income countries. 2021. DOI: https://www.nature.com/articles/d41586-021-02383-z.

12. Chookajorn, T., et al., Southeast Asia is an emerging hotspot for COVID-19. Nature Medicine, 2021. 27(9): p. 1495–1496.

13. Organization, W.H., COVID-19-WHO Thailand Situation Reports. 2022.

14. The Department of Disease Control, M.o.P.H., Thailand, COVID-19 Situation in Thailand. 2022.

15. OurWorldinData. Daily new cofirmed COVID-19 cases 2022; Available from: https://ourworldindata.org/explorers/coronavirus-data-explorer?zoomToSelection=true&time=2020-03-01..latest&facet=none&pickerSort=desc&pickerMetric=new_cases_smoothed_per_million&Metric=Confirmed+cases&Interval=7-day+rolling+average&Relative+to+Population=true&Color+by+test+positivity=false&country=~THA.

16. Vantanaprason, P. The Researcher COVID data. 2022; Available from: https://github.com/porames/the-researcher-covid-tracker.

17. OurWorldinData. COVID-19 vaccine doses administered. 2022; Available from: https://ourworldindata.org/grapher/cumulative-covid-vaccinations?country=~THA.

18. Prem, K., A.R. Cook, and M. Jit, Projecting social contact matrices in 152 countries using contact surveys and demographic data. PLoS Comput Biol, 2017. 13(9): p. e1005697.

19. Levin, A.T., et al., Assessing the age specificity of infection fatality rates for COVID-19: systematic review, meta-analysis, and public policy implications. Eur J Epidemiol, 2020. 35(12): p. 1123–1138.

20. Bubar, K.M., et al., Model-informed COVID-19 vaccine prioritization strategies by age and serostatus. Science, 2021. 371(6532): p. 916–921.

21. Gazit, S., et al., SARS-CoV-2 Naturally Acquired Immunity vs. Vaccine-induced Immunity, Reinfections versus Breakthrough Infections: a Retrospective Cohort Study. Clin Infect Dis, 2022.

22. Diekmann, O. and J. Heesterbeek, Mathematical Epidemiology of Infectious Diseases: Model Building, Analysis, and Interpretation. 2000: Wiley.

23. Davies, N.G., et al., Age-dependent effects in the transmission and control of COVID-19 epidemics. Nat Med, 2020. 26(8): p. 1205–1211.

24. Ferretti, L., et al., The timing of COVID-19 transmission. medRxiv, 2020: p. 2020.09.04.20188516.

25. Byambasuren, O., et al., Estimating the extent of asymptomatic COVID-19 and its potential for community transmission: systematic review and meta-analysis. medRxiv, 2020: p. 2020.05.10.20097543.

26. Eyre, D.W., et al., The impact of SARS-CoV-2 vaccination on Alpha & Delta variant transmission. medRxiv, 2021: p. 2021.09.28.21264260.

27. Lopez Bernal, J., et al., Effectiveness of Covid-19 Vaccines against the B.1.617.2 (Delta) Variant. New England Journal of Medicine, 2021. 385(7): p. 585–594.

28. Jara, A., et al., Effectiveness of an Inactivated SARS-CoV-2 Vaccine in Chile. New England Journal of Medicine, 2021. 385(10): p. 875–884.

29. Voysey, M., et al., Safety and efficacy of the ChAdOx1 nCoV-19 vaccine (AZD1222) against SARS-CoV-2: an interim analysis of four randomised controlled trials in Brazil, South Africa, and the UK. Lancet, 2021. 397(10269): p. 99–111.

30. Moghadas, S.M., et al., Evaluation of COVID-19 vaccination strategies with a delayed second dose. PLoS Biol, 2021. 19(4): p. e3001211.

31. Organization, W.H. Evidence Assessment: Sinovac/CoronaVac COVID-19 vaccine. 2021, Arpil Available from: https://cdn.who.int/media/docs/default-source/immunization/sage/2021/april/5_sage29apr2021_critical-evidence_sinovac.pdf.

32. Panarat Thepgumpanat, P.W.-u. Thailand considering limits on AstraZeneca vaccine exports. 2021; Available from: https://www.reuters.com/world/asia-pacific/thailand-considering-regulating-volume-astrazeneca-vaccine-exports-health-2021-07-14/.

33. Wanlapakorn, N., et al., Safety and immunogenicity of heterologous and homologous inactivated and adenoviral-vectored COVID-19 vaccines in healthy adults. medRxiv, 2021: p. 2021.11.04.21265908.

34. Thailand, C.f.C.-S.A.C.o., Thailand situation. 2020.

35. Mak, H.-Y.a.D., Tinglong and Tang, Christopher S., Managing Two-Dose COVID-19 Vaccine Rollouts with Limited Supply: Operations Strategies for Distributing Time-Sensitive Resources (August 28, 2021). SSRN, 2021.

36. Wilasang, C., et al., Reconstruction of the transmission dynamics of the first COVID-19 epidemic wave in Thailand. Scientific Reports, 2022. 12(1): p. 2002.

37. Reuters Royal academy seek 1 mln vaccines as Thailand approves Sinopharm. 2021.

38. Cori, A., et al., A new framework and software to estimate time-varying reproduction numbers during epidemics. Am J Epidemiol, 2013. 178(9): p. 1505–12.

39. Reuters. Thailand expands lockdown areas as COVID-19 cases surge. 2021 [cited 2021 18 November]; Available from: https://www.reuters.com/world/asia-pacific/thailand-expands-lockdown-areas-covid-19-cases-surge-2021-07-18/.

40. Mathieu, E., et al., Author Correction: A global database of COVID-19 vaccinations. Nat Hum Behav, 2021. 5(7): p. 956–959.

41. OurWorldinData. Available from: https://ourworldindata.org/grapher/covax-donations?country=FRA~ESP~SWE~USA~CAN~NOR~NZL~GBR~DNK~CHE~ITA~DEU~PRT~ARE~BEL~European+Union~JPN~NLD~FIN~HKG~IRL.

42. Organization, W.H. World Health Organization. Update on Omicron. 2021; Available from: https://www.who.int/news/item/28-11-2021-update-on-omicron.

43. OurWorldinData. Live COVID-19 vaccination tracker. 2022; Available from: https://covidvax.live/location/tha.

44. Panarat Thepgumpanat, P.W.-u. In first, Thailand to mix Sinovac, AstraZeneca vaccine doses. 2021; Available from: https://www.reuters.com/world/asia-pacific/thailand-starts-tighter-coronavirus-lockdown-around-capital-2021-07-12/.

45. WHO. Interim recommendations for use of the ChAdOx1-S [recombinant] vaccine against COVID-19 (AstraZeneca COVID-19 vaccine AZD1222 VaxzevriaTM, SII COVISHIELDTM). 2021; Available from: https://www.who.int/publications/i/item/WHO-2019-nCoV-vaccines-SAGE_recommendation-AZD1222-2021.1.

46. Emary, K.R., et al., Efficacy of ChAdOx1 nCoV-19 (AZD1222) vaccine against SARS-CoV-2 variant of concern 202012/01 (B. 1.1. 7): an exploratory analysis of a randomised controlled trial. The Lancet, 2021. 397(10282): p. 1351–1362.

47. Cerqueira-Silva, T., et al., Vaccine effectiveness of heterologous CoronaVac plus BNT162b2 in Brazil. Nature Medicine, 2022: p. 1–6.

48. Akpolat, T. and O. Uzun, Reduced mortality rate after coronavac vaccine among healthcare workers. Journal of Infection, 2021. 83(2): p. e20–e21.

49. Excler, J.-L., et al., Vaccine development for emerging infectious diseases. Nature medicine, 2021. 27(4): p. 591–600.

50. Wilasang, C., et al., Reduction in effective reproduction number of COVID-19 is higher in countries employing active case detection with prompt isolation. Journal of travel medicine, 2020. 27(5): p. taaa095.

51. Minoza, J.M.A., V.P. Bongolan, and J.F. Rayo, COVID-19 Agent-Based Model with Multi-objective Optimization for Vaccine Distribution. arXiv preprint arXiv:2101.11400, 2021.

52. Buckner, J.H., G. Chowell, and M.R. Springborn, Dynamic prioritization of COVID-19 vaccines when social distancing is limited for essential workers. Proceedings of the National Academy of Sciences, 2021. 118(16): p. e2025786118.

53. Chen, J., et al., Prioritizing allocation of COVID-19 vaccines based on social contacts increases vaccination effectiveness. medRxiv, 2021.

54. Shim, E., Optimal Allocation of the Limited COVID-19 Vaccine Supply in South Korea. Journal of Clinical Medicine, 2021. 10(4): p. 591.

## References

1. Bubar, K.M., et al., Model-informed COVID-19 vaccine prioritization strategies by age and serostatus. Science, 2021. 371(6532): p. 916–921.

2. Mathieu, E., et al., Author Correction: A global database of COVID-19 vaccinations. Nat Hum Behav, 2021. 5(7): p. 956–959.

